# Longer Sleep Duration Predicts Progression to Bipolar or Psychotic Disorders in Youth accessing Early Intervention Mental Health Services

**DOI:** 10.64898/2026.03.04.26347669

**Authors:** Joanne S. Carpenter, Jacob J. Crouse, Mathew Varidel, Emiliana Tonini, Mirim Shin, Natalia Zmicerevska, Alissa Nichles, Daniel F Hermens, Kathleen R Merikangas, Elizabeth M. Scott, Ian B. Hickie, Frank Iorfino

**Affiliations:** Youth Mental Health and Technology Team, Brain and Mind Centre, The University of Sydney, NSW Australia; Thompson Institute, University of the Sunshine Coast, Birtinya, Australia; Genetic Epidemiology Branch, Intramural Research Program, National Institute of Mental Health, Bethesda, USA

**Keywords:** Actigraphy, mental health, early intervention, risk prediction, youth, bipolar disorder, psychotic disorders, sleep, circadian

## Abstract

**Background:** While growing evidence implicates sleep-wake and circadian rhythm disturbances (SCRDs) in the onset and course of mood and psychotic disorders, longitudinal studies using objective measures are limited. This clinical cohort study examined whether actigraphy-derived SCRDs (sleep duration, timing, and efficiency) predicted transition to (i) any full-threshold mental disorders; and then specifically: (ii) full-threshold bipolar or psychotic disorders or (iii) other full-threshold (i.e. depressive or anxiety) disorders, in youth accessing mental health care.

**Methods:** Actigraphy monitoring was completed for 5-23 days in 250 participants (aged 12-30) presenting to youth-focused early intervention services in Sydney, Australia. Participants were followed longitudinally as part of the Optymise cohort for 6+ months (up to 8 years; median 2.5 years). Logistic regression and Cox proportional hazard models estimated associations between SCRDs and illness progression, after controlling for relevant baseline clinical and demographic covariates (e.g., age, sex, social and occupational functioning, mania-like and psychotic-like experiences, medication use).

**Results:** Longer sleep duration at baseline predicted higher odds of transition (OR = 2.23 [95%CI = 1.38-3.74]), and shorter time-to-transition (HR = 2.05 [95%CI = 1.23-3.40]) to full-threshold bipolar or psychotic disorders. This effect remained significant after controlling for clinical covariates. Later sleep midpoint predicted transition to any full-threshold mental disorder (OR = 1.46 [95%CI = 1.02-2.17]) at the uncorrected significance level.

**Conclusions:** Excessive sleep duration may represent an early marker of vulnerability for progression to severe mental illness. Findings support the prognostic utility of objective measures of SCRDs to guide indicated prevention and early intervention.

## INTRODUCTION

Sleep-wake and circadian disturbances (SCRDs) are frequently observed in mood and psychotic disorders and have been associated with both illness onset and course. Cross-sectional studies demonstrate altered sleep duration, timing, and variability across a range of mental health diagnoses, with circadian disruption increasingly recognised as a potential pathophysiological mechanism underlying a subset of cases ^1,2^, and highlighted as a key priority research area ^3^. Longitudinally, there is evidence that subjective SCRDs have bidirectional associations with mental disorders ^4-6^, with poor sleep quality, insomnia, hypersomnia, delayed timing, and greater day-to-day variability in sleep-wake measures predicting both onset of illness episodes and recurrence ^7-13^. Despite this evidence, longitudinal studies using objective measures of sleep-wake and circadian function as predictors of clinical outcomes remain very sparse.

Actigraphy (or accelerometry) monitoring offers a practical, scalable, and ecologically valid approach to objectively measuring sleep-wake and circadian behaviour, providing a robust picture of rest and activity patterns over multiple days ^14^. In clinical populations with established mental disorders, actigraphy studies generally report that later timing and more variable sleep are associated with relapse or recurrence of both depressive and manic/hypomanic episodes ^15-17^. In community-based samples, shorter objective sleep duration, and less robust or dampened rest-activity rhythms have been associated with increased depressive or anxiety symptoms ^18,19^ and increased incidence of major depressive or anxiety disorders across longitudinal follow up ^20^. There is also some longitudinal evidence that actigraphy-measured shorter sleep duration, reduced sleep efficiency, reduced activity, and increased variability in activity patterns is associated with subsequent development of psychotic symptoms in those at ultra-high risk for psychosis ^21,22^. Finally, we have previously reported that reduced sleep efficiency predicts longitudinal worsening of manic symptoms in young people presenting for mental health care ^23^. Aside from this, there is very limited evidence for the predictive value of objective SCRD measures among transdiagnostic clinical populations in the early stages of illness, when prognostic markers-especially those distinguishing trajectories driven by SCRDs-may be most useful for informing early intervention strategies ^24,25^.

A critical clinical outcome in this context is transition to full-threshold mental disorders, particularly severe conditions such as bipolar or psychotic disorders. Transition to a full-threshold disorder carries significant prognostic implications, including greater illness burden, increased risk of chronicity, and poorer functional outcomes, especially when not detected early ^26,27^. There are major delays in time to diagnosis and treatment for bipolar and psychotic disorders^28,29^, highlighting an urgent need to identify useful prognostic markers early in the course of illness ^30^. Currently, subthreshold states represent some of the strongest predictors of transition to diagnosable disorders ^31,32^ however objective markers with predictive value would add considerable clinical utility in the early stages of illness, and may also be informative prior to the development of subthreshold symptoms.

Cross-sectionally across the psychosis spectrum, individuals at clinical high risk exhibit reduced sleep efficiency, whereas those in the early stages of illness tend to show longer sleep duration ^33^. For bipolar disorders, later sleep-wake timing and longer sleep duration appear to distinguish those with recent onset disorders from healthy controls ^34^. However, effects in at-risk populations are less clear. For example, offspring of individuals with bipolar disorder show longer sleep and later wake times, whereas individuals with subthreshold symptom manifestations demonstrate shorter sleep duration and increased variability ^34^. In both bipolar and psychotic illness, medication use has been identified as a likely confounding factor, particularly in relation to sleep duration findings ^33,34^.

Accordingly, the aim of the present study was to examine the longitudinal associations between actigraphy-derived SCRDs (sleep timing, duration, efficiency, and variability) measured around the time of entry to early intervention services and longitudinal risk of transition to (i) any full-threshold mental disorders; (ii) full-threshold bipolar or psychotic disorders; and (iii) other full-threshold disorders (i.e. unipolar depressive and anxiety disorders) in the Optymise cohort - a sample of young people presenting to early intervention mental health services in Sydney, Australia ^35^. We have previously reported on associations between clinical features and longitudinal illness outcomes more broadly in this cohort ^36,37^; and more specific research on the longitudinal association between sleep-wake profiles and subsequent manic symptoms and memory impairments ^23^ as well as cross-sectional studies focussing on actigraphy features across diagnostic categories ^38,39^. Here, we focus on a subsample of the Optymise cohort with available actigraphy data at baseline and detailed longitudinal follow up across the course of care (at least 1 follow up timepoint 6+ months from baseline). Based on previous (but largely cross-sectional and/or subjective) sleep-wake research, we hypothesised that increased risk for transitions to full-threshold disorders and onset of at-risk mental states would be associated with both short and long sleep duration, later sleep timing, poorer sleep efficiency, and more variable sleep duration and timing. These analyses and hypotheses were not pre-registered.

## METHODS

### Participants

Participants were drawn from the Brain and Mind Centre’s Optymise cohort. This cohort includes 6743 individuals aged 12-30 years old who presented to the Brain and Mind Centre’s early intervention youth mental health clinics (in Sydney, Australia) and were recruited to a clinical research register between June 2008 and July 2018 ^35^. Clinical longitudinal data was available for 2901 individuals. A subset of this cohort completed actigraphy monitoring as part of a linked in-depth neurobiological sub-study: the Youth Mental Health Follow Up Study ^40^. Participants were excluded for insufficient English language skills to comprehend the research protocol, clinically evident intellectual disability, history of neurological disease (e.g. epilepsy), medical illness known to impact cognitive and brain function (e.g. cancer), electroconvulsive therapy in the last 3 months, and/or current substance dependence.

Participants were selected for the current analysis if they met the following criteria (i) completion of actigraphy monitoring with at least 5 days of valid actigraphy data; and (ii) at least 6 months of clinical research follow up the from time of actigraphy recording. A flow diagram of inclusion and exclusion is included in the supplementary materials (Figure S1). The University of Sydney Human Research Ethics Committee approved this research (project numbers 2008/5453, 2012/1626, and 2012/1631). Participants (or their guardians if under 16 years of age) provided written informed consent.

### Data Collection

A detailed description of data collection for the Optymise cohort is provided in a cohort profile ^35^. Briefly, data on specific illness course characteristics were extracted from clinical and research files at predetermined time points and entered into a standardized clinical proforma. The first available clinical assessment at the mental health service was taken as the baseline time point for each participant and the date of this assessment was used to determine each of the follow-up time points: 3 months, 6 months, 12 months, 2 years, 3 years, 4 years and 5 years. If there was no clinical information available for any time point (i.e. the participant did not attend the service during that time) then that entry was left missing. A ‘time last seen’ (TLS) entry was also used to capture clinical information from the most recent presentation to the clinical service, which did not always align with one of the pre-specified time points. All clinical and research notes from the preceding time points, up to and including the current time point were used to inform and complete the current proforma entry.

In the present study, relevant measures from the proforma include: current diagnosis (based on Diagnostic and Statistical Manual of Mental Disorders [DSM-5 ^41^] criteria); demographic features (e.g., age, sex); social and occupational function (measured by the Social and Occupational Functioning Assessment Scale [SOFAS ^42^], at-risk mental states, and treatment utilization. At-risk mental states included psychotic-like experiences—which were defined as the presence of any psychotic symptoms including: perceptual abnormalities, bizarre ideas, disorganised speech, psychotic-like unusual language or thought content, or psychotic-like disruptive or aggressive behaviour—and manic-like experiences, which were defined as the presence of any manic/hypomanic symptoms including: abnormally elevated mood or irritability; increased motor activity, speech, or sexual interest; manic-like disruptive or aggressive behaviour; manic-like unusual language or thought content; increased goal directed behaviour; or decreased need for sleep. SOFAS was missing for 5 participants.

For the purposes of this analysis, the Optymise time point closest to the available actigraphy recording was used as the baseline time point. For the actigraphy component of the study, participants were provided with a wrist-worn actigraphy recording devices (Actiwatch-64/L/2, Philips Respironics, Pittsburgh, USA; or GENEActiv, Activinsights, Kimbolton, UK) for between 5 to 23 days (median 13 days) in their usual home environment (Note: a protocol change during the Youth Mental Health Research Study prompted a change from the Actiwatch to the GENEActiv device). Participants were instructed to wear the device on their non-dominant wrist and only remove it for showering, bathing, or swimming. Actiwatch data was collected over 30-second or 1-minute epochs; GENEActiv data was collected at 50hz. Where more than one actigraphy recording was available for the same participant, the earliest recording was used.

### Actigraphy Data Processing

As the two types of actigraphy devices used (Actiwatch, GENEActiv) provide different raw motor activity metrics (activity count [AC] and Euclidean Norm Minus One [ENMO], respectively), processing was completed using two different pipelines.

Raw GENEActiv actigraphy data was processed using an open-source R package, GGIR (version 3.2.6), developed to process multi-day accelerometer data ^43^. Post-processing was completed using the package mMARCH.AC (version 2.9.4) according to protocols established for the National Institute of Mental Health’s Mobile Motor Activity Research Consortium for Health (mMARCH) ^44^. Any days with less than 8 hours of data were removed. Missing data on remaining days was imputed with the mean of that epoch on all other days. GGIR and mMARCH.AC algorithms were used to generate sleep parameters using established algorithms ^45^.

Actiwatch data was visually inspected by trained technicians to adjust the start and end of each estimated sleep episode and identify exclusion periods where the watch was removed. Standard algorithms for Actiwatch data ^46^ were used to identify periods of wake during the sleep episode with a medium sensitivity threshold of 40 counts per epoch.

Of the final sample, 217 recordings were from Actiwatch devices and 37 were from GENEActiv devices. For both devices, the following actigraphy measures were generated for each day of recording: sleep duration (number of minutes estimated asleep per 24-hour period); sleep midpoint (the halfway point between the onset of sleep and wake time); and sleep efficiency (the percentage of time estimated as asleep between sleep onset and wake time). These variables were chosen as parameters that were available and comparable from the two different recording devices and processing pipelines. They also provide information about three key aspects of sleep-wake patterns: duration, timing, and quality. In order to quantify variability across the recording period in addition to average duration, timing and efficiency, both the mean and standard deviation were calculated for each of the three parameters for each participant. Two individuals (one Actiwatch device, one GENEActiv device) were excluded from analyses due to actigraphy patterns identified as potential Non-24-Hour Sleep-Wake Disorder (based on visual inspection and extreme standard deviation of sleep midpoint >4-hours).

### Statistical Analyses

Analyses were performed in R (version 4.4.2) and RStudio (version 2025.5.0). To minimize the influence of extreme outliers, all actigraphy variables were windsorized to ±3 standard deviations (less than 2% of values were transformed, with between 2 and 8 values windsorized per variable). Longitudinal outcomes were (i) the presence of any full-threshold mental disorder, including any of the following: depressive disorders, bipolar disorders, anxiety disorders, psychotic disorders, or obsessive compulsive disorder; (ii) the presence of a full-threshold bipolar or psychotic disorder including full-threshold bipolar I, bipolar II, or psychotic disorder; and (iii) the presence of other full-threshold disorders (i.e. those included in i but not ii). Initial examination of cross-sectional associations between these two clinical outcomes and each sleep-wake parameter at baseline was performed using logistic regression. Next, longitudinal associations between each sleep-wake parameter at baseline and transition to each of the clinical outcomes across follow up was performed using logistic regression (participants who already had the outcome at baseline were excluded from these follow-up analyses). For all logistic regression analyses, p-values were adjusted per outcome using the Bonferroni method (i.e. six tests per outcome for the six sleep-wake parameters). Finally, time-to-transition to each longitudinal outcome was analysed using Cox proportional hazards models including all six sleep-wake parameters as simultaneously-modelled predictors. Assigned sex at birth and baseline age were included as covariates for all analyses. A p-value of <.05 was considered statistically significant for all analyses.

As post-hoc analyses for any significant associations, a second model (i.e. logistic regression and/or Cox proportional hazards) was fitted including additional covariates selected based on prior evidence of their influence on sleep-wake parameters and/or each outcome, or on the basis of significant univariate associations (see supplementary figure S2). These included SOFAS, NEET status (not in education, employment or training), mania-like experiences, psychotic-like experiences, and medication use (antidepressant, antipsychotic, or mood stabilizer use). As supplementary analyses, any significant findings relating to outcome (ii) (i.e. transition to full-threshold bipolar or psychotic disorder) were explored in separate models for (a) bipolar disorders (I or II) and (b) psychotic disorders. Given the low rates of transition to these specific outcomes and associated low power, these analyses were considered exploratory. Given the hypothesized effects of both long and short sleep duration and potential effects of both early and late sleep timing, additional supplementary analyses included both linear and quadratic terms for sleep duration and sleep midpoint as predictors.

## RESULTS

There were 254 participants who met our eligibility criteria and were included in the final analysis (64% female; mean age ± SD = 19.9 ± 3.9 years). The duration of follow-up since the actigraphy recording ranged from 6 months to 8 years (median = 2.5 years; IQR = 2.4 years) with a median of 4 follow-up assessments (range 1-8 follow-up assessments). Table 1 reports baseline characteristics of the included actigraphy sample and the full Optymise Cohort with available clinical data. The included actigraphy sample were older at baseline, had lower SOFAS, were more likely to have mania-like experiences, more likely to have a full threshold disorder (specifically depressive, bipolar, or anxiety disorders), and were more likely to be taking psychotropic medication (antidepressant, antipsychotic or mood stabilizers). Rates of transition to specified outcomes are reported in Table 2.

**Table 1.**
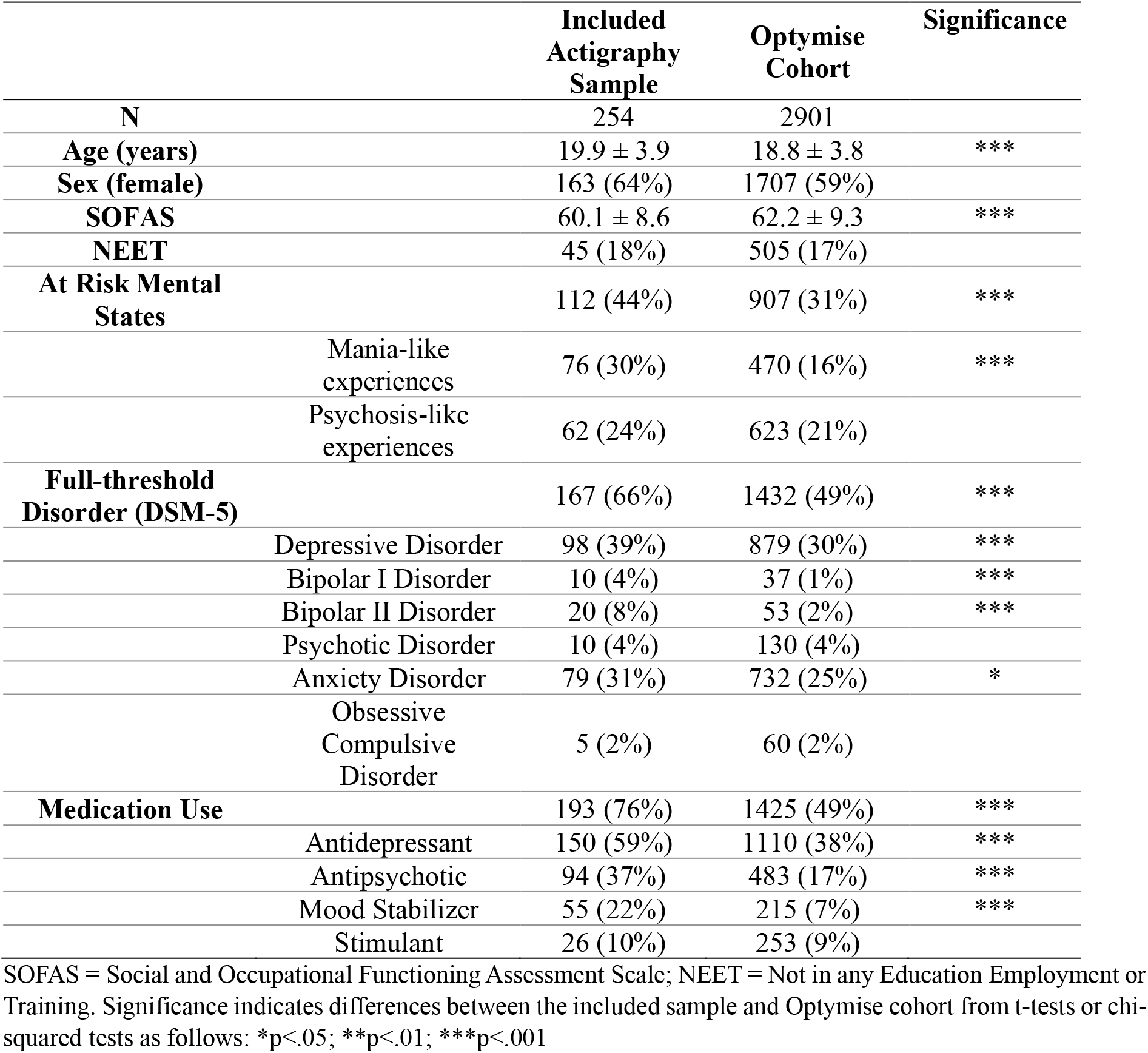
Characteristics of the included sample and the full Optymise Cohort at baseline.

|  |  | Included<br>Actigraphy<br>Sample | Optymise<br>Cohort | Significance |
| --- | --- | --- | --- | --- |
| N |  | 254 | 2901 |  |
| Age (years) |  | 19.9 ± 3.9 | 18.8 ± 3.8 | *** |
| Sex (female) |  | 163 (64%) | 1707 (59%) |  |
| SOFAS |  | 60.1 ± 8.6 | 62.2 ± 9.3 | *** |
| NEET |  | 45 (18%) | 505 (17%) |  |
| At Risk Mental<br>States |  | 112 (44%) | 907 (31%) | *** |
|  | Mania-like<br>experiences | 76 (30%) | 470 (16%) | *** |
|  | Psychosis-like<br>experiences | 62 (24%) | 623 (21%) |  |
| Full-threshold<br>Disorder (DSM-5) |  | 167 (66%) | 1432 (49%) | *** |
|  | Depressive Disorder | 98 (39%) | 879 (30%) | *** |
|  | Bipolar I Disorder | 10 (4%) | 37 (1%) | *** |
|  | Bipolar II Disorder | 20 (8%) | 53 (2%) | *** |
|  | Psychotic Disorder | 10 (4%) | 130 (4%) |  |
|  | Anxiety Disorder | 79 (31%) | 732 (25%) | * |
|  | Obsessive<br>Compulsive<br>Disorder | 5 (2%) | 60 (2%) |  |
| Medication Use |  | 193 (76%) | 1425 (49%) | *** |
|  | Antidepressant | 150 (59%) | 1110 (38%) | *** |
|  | Antipsychotic | 94 (37%) | 483 (17%) | *** |
|  | Mood Stabilizer | 55 (22%) | 215 (7%) | *** |
|  | Stimulant | 26 (10%) | 253 (9%) |  |

SOFAS = Social and Occupational Functioning Assessment Scale; NEET = Not in any Education Employment or Training. Significance indicates differences between the included sample and Optymise cohort from t-tests or chi-squared tests as follows: \*p<.05; \*\*p<.01; \*\*\*p<.001

**Table 2.** Transition rates to clinical outcomes across follow-up.

|  | **Transitioned over**<br>**follow up** | **Did not transition**<br>**by time last seen** | **Time to transition**<br>**(days)**<br>**(median[IQR])** |
| --- | --- | --- | --- |
| **Any Full-threshold**<br>**Disorder** | 31 (36%) | 56 (64%) | 269 [683] |
| **Full-threshold Bipolar or Psychotic Disorder** | 22 (10%) | 192 (90%) | 592 [773] |
| **Other Full-threshold Disorder (Depressive or Anxious Disorders)** | 23 (29%) | 56 (71%)<sup>a</sup> | 318 [823] |

<sup>a</sup>excludes those that transitioned to Full-threshold Bipolar or Psychotic Disorder

### SCRDs and baseline full-threshold disorders

Logistic regression models quantifying cross-sectional associations between sleep-wake measures and clinical characteristics at baseline are reported in Table 3. First, there were no associations between the presence of any full-threshold disorder at baseline and SCRDs. Second, in those with full-threshold bipolar or psychotic disorders at baseline, higher sleep efficiency was a significant predictor (OR = 1.14; 95% CI = 1.05-1.25; p = .004) corresponding to an absolute risk increase from 15.7% to approximately 17.5% for a 1 percentage increase in sleep efficiency. Post-hoc analyses indicated that this effect was attenuated and no longer statistically significant when controlling for baseline social and occupational functioning, NEET status, presence of mania-like or psychotic-like experiences and mood stabilizer use (OR = 1.10; 95% CI = 1.00-1.23; p = .063). Supplementary analyses further indicated that this association was largely driven by higher sleep efficiency in those with full-threshold psychotic disorders (Table S1). Lower variation (SD) in sleep efficiency was also associated with the presence of a full-threshold bipolar or psychotic disorder at baseline (OR = 0.80; 95% CI = 0.64-0.98; p = .044), however this was no longer statistically significant following correction for multiple comparisons. There were no significant associations between any clinical characteristic and the mean (or SD) of sleep midpoint. There were no significant associations between the presence of any other (unipolar depressive or anxious) full-threshold disorder and any SCRDs.

**Table 3.** Associations between sleep-wake measures and clinical characteristics at baseline.

| Characteristic | Predictors | Odds Ratio<br>(95% CI) | P value | Adjusted<br>p value | Covariates |
| --- | --- | --- | --- | --- | --- |
| **Any Full-threshold Disorder** | Sleep Midpoint | 1.04 (0.85-1.27) | 0.696 | 1 | Age, Sex |
|  | Sleep Duration | 1.21 (0.91-1.61) | 0.192 | 1 | Age, Sex |
|  | Sleep Efficiency | 1.04 (0.99-1.10) | 0.145 | 0.872 | Age, Sex |
|  | SD of Midpoint | 1.10 (0.71-1.75) | 0.684 | 1 | Age, Sex |
|  | SD of Duration | 1.06 (0.63-1.81) | 0.821 | 1 | Age, Sex |
|  | SD of Efficiency | 0.98 (0.85-1.13) | 0.815 | 1 | Age, Sex |
| **Full-threshold Bipolar or Psychotic Disorder** | Sleep Midpoint | 0.98 (0.76-1.25) | 0.883 | 1 | Age, Sex |
|  | Sleep Duration | 1.35 (0.97-1.90) | 0.078 | 0.469 | Age, Sex |
|  | Sleep Efficiency | 1.14 (1.05-1.25) | 0.004** | 0.026* | Age, Sex |
|  | Sleep Efficiency<sup>†</sup> | 1.10 (1.00-1.23) | 0.063 | - | Age, Sex, MLE, PLE, SOFAS, NEET, Mood Stabiliser Use |
|  | SD of Midpoint | 0.72 (0.37-1.28) | 0.289 | 1 | Age, Sex |
|  | SD of Duration | 0.76 (0.37-1.50) | 0.437 | 1 | Age, Sex |
|  | SD of Efficiency | 0.80 (0.64-0.98) | 0.044* | 0.263 | Age, Sex |
| **Other Full-threshold Disorder (Depressive or Anxious Disorder)** |  |  |  |  |  |
|  | Sleep Midpoint | 1.09 (0.90-1.31) | 0.386 | 1 | Age, Sex |
|  | Sleep Duration | 1.04 (0.81-1.35) | 0.755 | 1 | Age, Sex |
|  | Sleep Efficiency | 1.00 (0.94-1.05) | 0.885 | 1 | Age, Sex |
|  | SD of Midpoint | 1.36 (0.89-2.11) | 0.156 | 0.938 | Age, Sex |
|  | SD of Duration | 1.31 (0.80-2.17) | 0.278 | 1 | Age, Sex |
|  | SD of Efficiency | 1.07 (0.93-1.22) | 0.338 | 1 | Age, Sex |

Results presented are logistic regression models adjusted for listed covariates. Adjusted p values are Bonferroni corrected per outcome variable. CI = Confidence interval. MLE = Mania-like experiences. PLE = Psychosis-like experiences. \*p<.05; \*\*p<.01; \*\*\*p<.001. † indicates post-hoc analysis.

### SCRDs and longitudinal clinical outcomes

Logistic regression models predicting clinical outcomes across longitudinal follow-up from sleep-wake measures are reported in Table 4. Later sleep midpoint was associated with increased odds of developing any full-threshold disorder across follow-up (OR = 1.46; 95% CI = 1.02-2.17; p = .046), however this association was no longer significant following correction for multiple comparisons. Longer sleep duration was significantly associated with an increased odds of developing a full-threshold bipolar or psychotic disorder across follow up (OR = 2.23; 95% CI = 1.38-3.74; p = .001) corresponding to an absolute risk increase from 10% to approximately 19.9% for each additional hour of sleep duration. Post-hoc analyses indicated this effect remained significant when controlling for baseline social and occupational functioning, NEET status, presence of mania-like or psychotic-like experiences, mood stabilizer use, antipsychotic use, and antidepressant use (OR = 2.14; 95% CI = 1.26-3.79; p = .006). Supplementary analyses indicated that this effect was driven by both bipolar and psychotic disorders, with a stronger effect for psychotic disorders (Table S2). Group comparisons for significant associations at baseline or across follow-up are shown in supplementary Figure S3. There were no significant associations between any SCRDs and the odds of developing any other (unipolar depressive or anxious) full-threshold disorder.

**Table 4.** Associations between sleep-wake measures and clinical outcomes across longitudinal follow-up.

| Outcome | Predictors | Odds Ratio<br>(95% CI) | P value | Adjusted<br>p value | Covariates |
| --- | --- | --- | --- | --- | --- |
| **Any Full-threshold Disorder** | Sleep Midpoint | 1.46 (1.02-2.17) | 0.046* | 0.274 | Age, Sex |
|  | Sleep Duration | 1.11 (0.63-1.98) | 0.728 | 1 | Age, Sex |
|  | Sleep Efficiency | 1.02 (0.91-1.15) | 0.714 | 1 | Age, Sex |
|  | SD of Midpoint | 0.64 (0.21-1.68) | 0.393 | 1 | Age, Sex |
|  | SD of Duration | 1.14 (0.42-3.06) | 0.787 | 1 | Age, Sex |
|  | SD of Efficiency | 1.04 (0.81-1.34) | 0.742 | 1 | Age, Sex |
| **Full-threshold Bipolar or Psychotic Disorder** | Sleep Midpoint | 1.05 (0.75-1.45) | 0.788 | 1 | Age, Sex |
|  | Sleep Duration | 2.23 (1.38-3.74) | 0.001** | 0.009** | Age, Sex |
|  | Sleep Duration<sup>†</sup> | 2.14 (1.26-3.79) | 0.006** | - | Age, Sex, MLE, PLE, SOFAS, NEET, Mood Stabilizer Use, Antipsychotic Use, Antidepressant Use |
|  | Sleep Efficiency | 1.07 (0.96-1.20) | 0.235 | 1 | Age, Sex |
|  | SD of Midpoint | 0.63 (0.24-1.42) | 0.311 | 1 | Age, Sex |
|  | SD of Duration | 0.94 (0.37-2.25) | 0.885 | 1 | Age, Sex |
|  | SD of Efficiency | 0.85 (0.64-1.10) | 0.250 | 1 | Age, Sex |
| **Other Full-threshold Disorder (Depressive or Anxious Disorder)** | Sleep Midpoint | 1.30 (0.88-1.98) | 0.200 | 1 | Age, Sex |
|  | Sleep Duration | 0.87 (0.45-1.70) | 0.688 | 1 | Age, Sex |
|  | Sleep Efficiency | 1.00 (0.88-1.13) | 0.975 | 1 | Age, Sex |
|  | SD of Midpoint | 0.48 (0.13-1.43) | 0.231 | 1 | Age, Sex |
|  | SD of Duration | 1.07 (0.36-3.11) | 0.896 | 1 | Age, Sex |
|  | SD of Efficiency | 1.08 (0.82-1.41) | 0.566 | 1 | Age, Sex |

Results presented are logistic regression models adjusted for listed covariates. Adjusted p values are Bonferroni corrected per outcome variable. CI = Confidence interval. MLE = Mania-like experiences. PLE = Psychosis-like experiences. SOFAS = Social and Occupational Functioning Assessment Scale. NEET = Not in Education Employment or Training. \*p<.05; \*\*p<.01; \*\*\*p<.001. † indicates post-hoc analysis.

### SCRDs and time-to-transition to clinical outcomes

Associations between sleep-wake measures and time to transition to outcomes across follow-up are reported in Table 5 (Cox proportional hazard models). Longer sleep duration was associated with an increased hazard of transition to a full-threshold bipolar or psychotic disorder (HR = 2.05; 95% CI = 1.23-3.40; p = .006), indicating that for each additional hour of sleep individuals transition roughly twice as fast to bipolar or psychotic disorders. Post-hoc analyses indicated that this effect remained statistically significant when controlling for baseline SOFAS, NEET status, presence of mania-like or psychotic-like experiences, mood stabilizer use, antipsychotic use, and antidepressant use (HR = 1.91; 95% CI =1.11-3.27; p = .019).

**Table 5.**
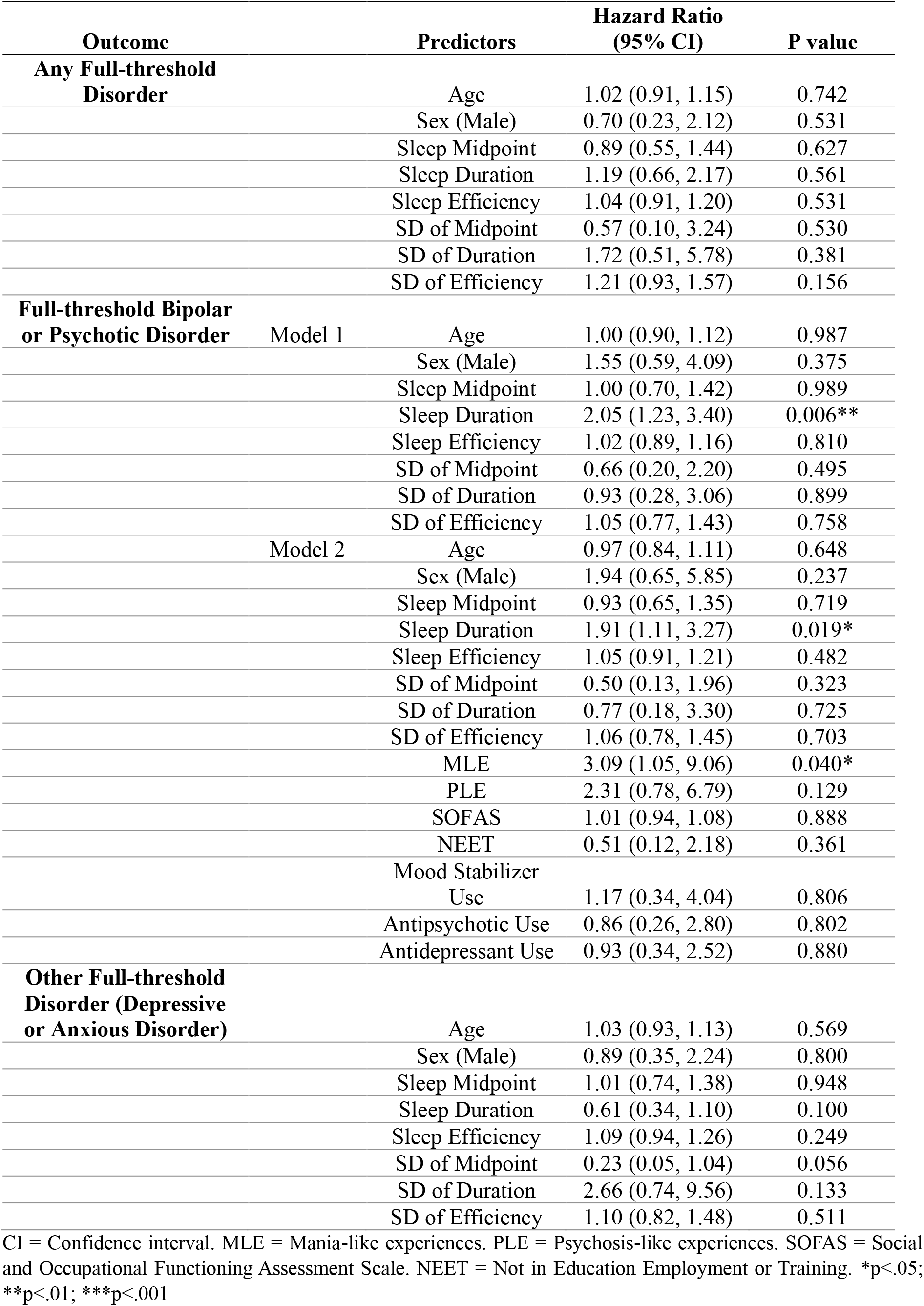
Cox proportional hazards models predicting time to transition to follow-up outcomes from sleep wake measures.

Figure 1 presents cumulative incidence curves for transition to a full-threshold bipolar or psychotic disorder based on sleep duration. For illustrative purposes, this figure presents groups defined by baseline sleep duration with cut points selected based on ±0.5 and ±1.5 SDs (all statistical analyses use continuous sleep parameters). Supplementary analyses indicated that the trend for effects of sleep duration on both odds of transition and time to transition was present in regard to both full-threshold bipolar and full-threshold psychotic disorders, with stronger effects for psychotic disorders (although the effect for time to transition to bipolar disorders did not reach significance; Table S3 and Figure S4).

**Figure 1.**
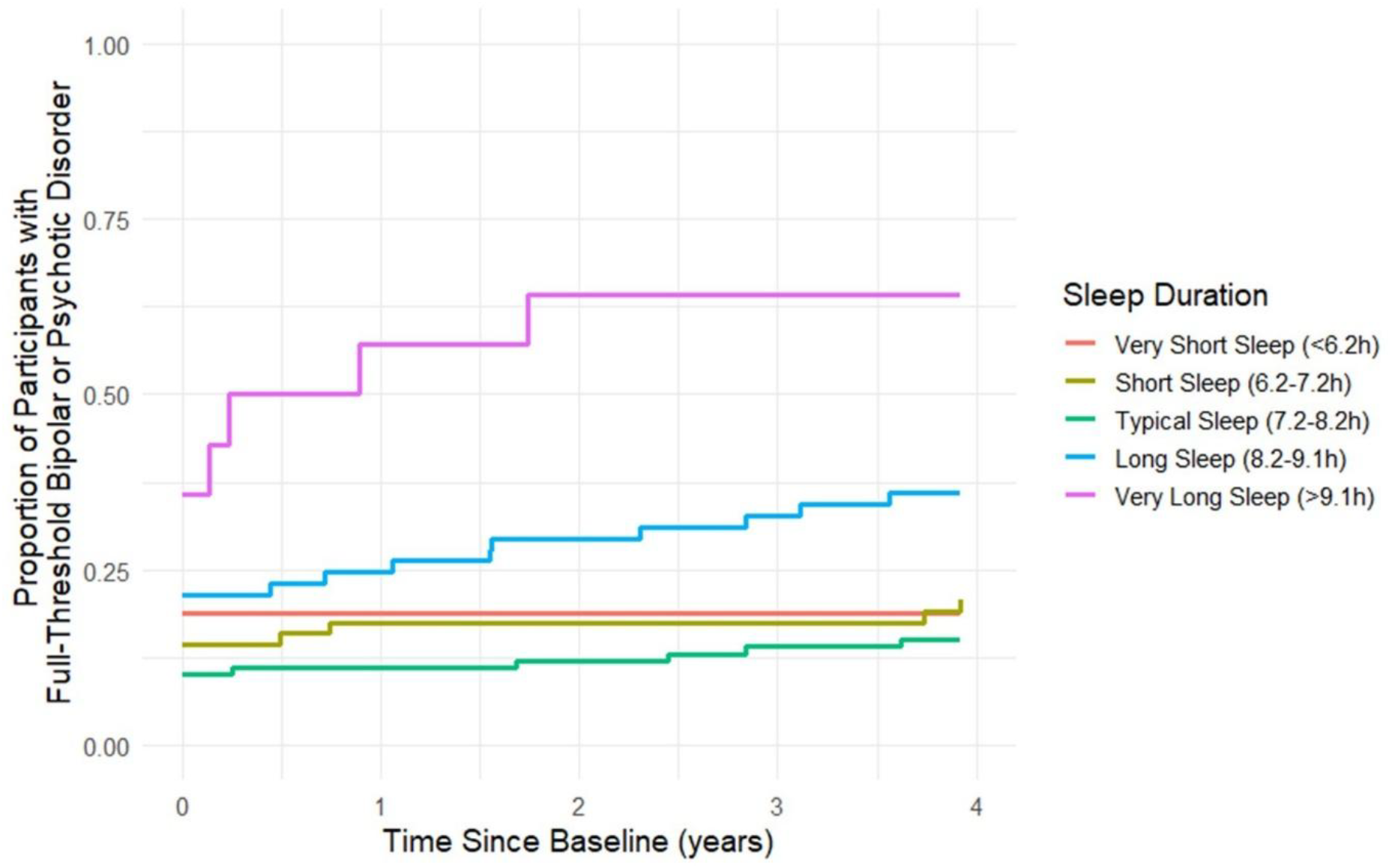
Cumulative Incidence of Transition to a Full-Threshold Bipolar or Psychotic Disorder by Sleep Duration. Sleep duration groups are defined for illustrative purposes based on mean ±0.5 and ±1.5 standard deviations.

Finally, supplementary analyses exploring quadratic terms for sleep duration and sleep midpoint across all analyses did not find significant quadratic effects, with the exception of an uncorrected quadratic effect of sleep duration on the presence of any full-threshold disorder at baseline (HR = 1.29; 95% CI = 1.01-1.66; p = .043) (Tables S4-S6, Figure S5).

## DISCUSSION

This study provides novel longitudinal evidence that objectively measured SCRDs are associated with illness progression in young people accessing early intervention mental health services in Sydney, Australia. The key finding was that longer sleep duration at baseline predicted both higher rates of transition and shorter time-to-transition to full-threshold bipolar or psychotic disorders. Additionally, later sleep midpoint predicted transition to any full-threshold disorder at the uncorrected significance level. By contrast, other hypothesised effects of variability in sleep timing and duration were not supported.

Our results highlight longer sleep duration as a potential early indicator of progression to more severe mental disorders (i.e. full-threshold bipolar or psychotic disorders) in young people presenting for care at transdiagnostic early intervention services. This builds on our previous cross-sectional findings in this population that longer sleep duration is associated with more advanced clinical stage ^39^ to suggest that excessive sleep duration may also be a prognostic marker. Importantly, the effect of sleep duration remained statistically significant when controlling for the presence of mania-like or psychosis-like experiences, use of psychotropic medications, social and occupational functioning, and NEET status. This strengthens the finding that long sleep duration may have predictive value above that of prodromal symptoms for predicting transition. The finding is in line with previous research reporting longer sleep duration in the early stages of bipolar and psychotic disorders and in those at familial risk ^33,34^, but contrasts with reports of shorter sleep duration in bipolar at-risk groups ^34^ and with longitudinal associations between shorter sleep duration and later psychotic symptoms ^22^. A potential explanation for these mixed findings is that both short and long sleep duration may signal increased mental disorder risk, potentially acting via different mechanisms. A significant quadratic effect was observed (at the uncorrected significance level: see supplementary table S4) for sleep duration on any baseline full-threshold disorder, and, inspection of Figure 1 (and Figure S4) suggests the possibility of a weaker (non-significant) association between short sleep and increased risk for transition to bipolar disorders in addition to the long sleep effect. The small transition numbers, and a potential high overall prevalence of insomnia (i.e. short sleep duration) in this clinical sample ^47^ may have reduced our ability to detect significant effects of short sleep in this study. The findings are partially consistent with our proposed transdiagnostic ‘circadian depression’ phenotype, which we hypothesise is characterised by hypersomnia, delayed sleep-wake (and circadian) timing, and increased variability in sleep-wake (and circadian) patterns ^1,25^. However, the present results, together with increasing evidence linking psychotic syndromes to circadian disturbance, suggest that this phenotype may not only be linked to bipolar risk, but instead span both bipolar and psychotic trajectories ^2^.

Contrary to our hypotheses, we did not observe significant predictive effects of variability in sleep duration or timing. One possibility to explain this is that these features may represent more state-based or proximal indicators of risk, while long sleep duration may reflect a more stable trait marker of vulnerability. It is also important to acknowledge methodological differences between this and previous research: our variability measures (SDs of duration, timing, and efficiency) may not capture the more nuanced circadian disruption reflected in other actigraphy metrics such as intra-daily variability, and inter-daily stability which have previously been linked to outcomes such as recurrence in bipolar disorder ^48,49^, but could not be calculated across the actigraphy devices used in the present study. Further, the short observation period (averaging two weeks) is not sufficient to capture long term stability and variability. Future research examining the effects of sleep-wake variability in larger samples and over longer observation periods will be important to more clearly understand associations with mental ill-health.

Several limitations warrant consideration. The focus on young people presenting to early intervention mental health services represents a population in which identification of risk is particularly important, however the findings may not translate into other settings such as community-based early identification and prevention programs. DSM-5 was retrospectively applied to the clinical information of this cohort, which limits comparability with other diagnostic methods. The cohort is also biased across follow-up to those who continue to engage in care, and the sample with available actigraphy were biased to include more full-threshold disorders (particularly depressive and bipolar disorders) and more medication use. As actigraphy was only available for a small proportion of the Optymise cohort, the transition rates to specific diagnoses and associated statistical power was reduced. Accordingly, alternative statistical methods used previously in this sample (e.g. multi-state modelling ^36,37^) were not appropriate. Further, due to the type of actigraphy devices used and harmonisation across different devices, it was not possible to examine alternative actigraphy metrics (e.g. intra-daily variability, or inter-daily stability, relative amplitude) that may better reflect SCRDs. While we did control for relevant classes of medication in our analyses, information about specific medications was not available, and so it was not possible to explore potential differential effects of specific medications e.g. typical vs atypical antipsychotics ^33^. It is also possible that there were confounding effects of other variables that were not controlled for such as body mass index, undiagnosed obstructive sleep apnoea, use of other sedating medications or substances (e.g., alcohol, cannabis). Finally, this study focused on baseline predictors of transition, however time-varying covariates, particularly with greater temporal resolution to the outcome, may provide additional insights.

Altogether, this study provides novel longitudinal evidence that objectively recorded SCRDs have prognostic value for severe mental illness outcomes in young people accessing early intervention services. Specifically, long sleep duration emerged as a robust predictor of transition to bipolar and psychotic disorders, even after accounting for prodromal symptoms and several known clinical confounders. This extends models of sleep-wake—and potentially underlying circadian disturbance (this is more speculative)—as a driver of vulnerability by highlighting that increased rather than reduced sleep duration may be particularly salient in some high-risk youth. These findings underscore the potential of objective sleep-wake and circadian assessment as a risk identification tool in early intervention settings, where they may inform directed intervention approaches that target SCRDs. Future work should aim to replicate these findings in larger, and community-based samples, integrate alternative circadian metrics, and adopt time-varying approaches to better capture dynamic risk processes. Identifying reliable and accessible markers of progression remains critical for refining early intervention strategies and improving outcomes for young people at risk of severe mood and psychotic disorders.

## Data Availability

All data produced in the present study are available upon reasonable request to the authors

## Author contributions

JSC, JJC, DFH, EMS, IBH, and FI designed the study. JSC, JJC, NZ, AN, and FI contributed to acquisition of data. JSC and MV conducted data analysis. JSC, JJC, ET, MS, KRM, JS, IBH and FI interpreted the analyses. All authors contributed to and approved the final manuscript.

## Acknowledgements

The authors acknowledge the Gadigal people of the Eora nation, upon whose ancestral lands our research was conducted. We pay our respect to elders past, present, and emerging. We thank all the young people who have participated in this study, and all the staff in the Youth Mental Health Team at the Brain and Mind Centre, past and present, who have contributed to this work.

