## Supplementary for "Longer Sleep Duration Predicts Progression to Bipolar or Psychotic Disorders in Youth accessing Early Intervention Mental Health Services"

Supplementary Information: Actigraphy-Measured Sleep Duration Predicts Transition to Full-Threshold Bipolar or Psychotic Disorders in Young People with Emerging Mood Disorders

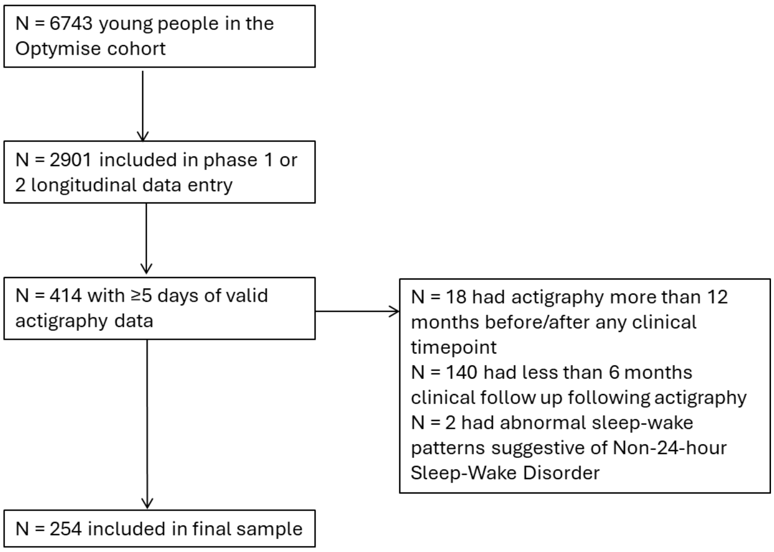


**Supplementary Figure S1.** Flow diagram of inclusion and exclusion from the Optymise Cohort

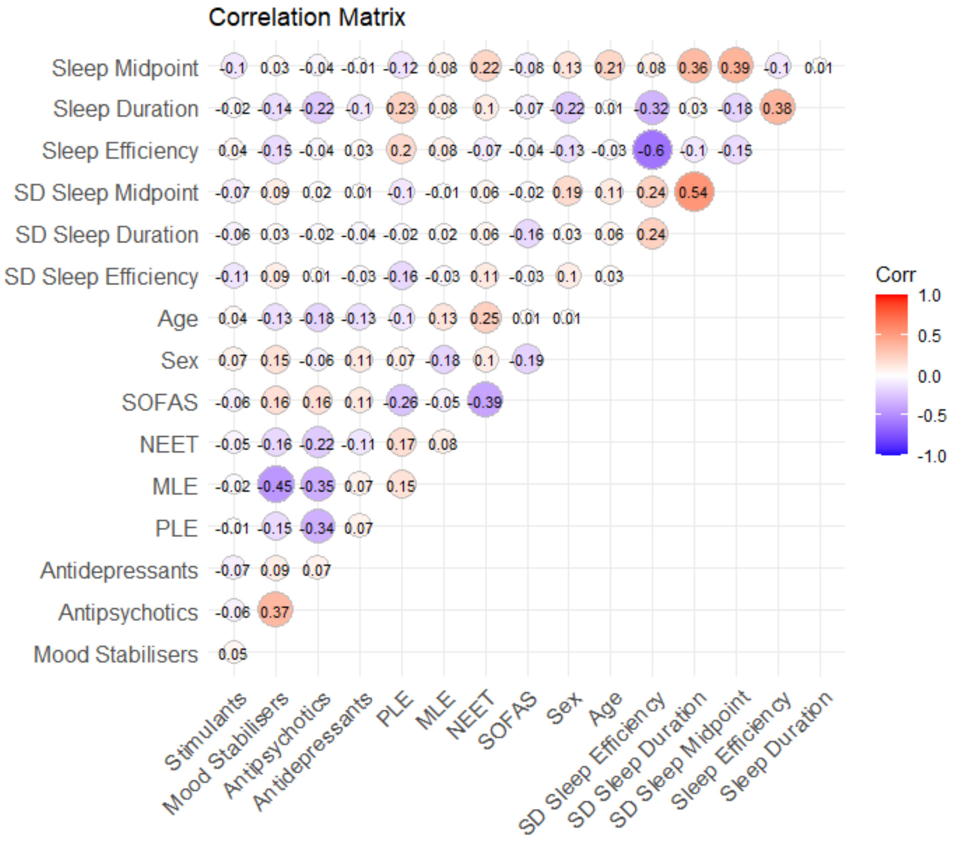


**Supplementary Figure S2.** Univariate associations between sleep-wake variables and other clinical covariates

**Supplementary Table S1.** Associations between sleep-wake measures and specific baseline outcomes

| Outcome | Predictors | Odds Ratio (95% CI) | P value | Adjusted p value | Covariates |
|---|---|---|---|---|---|
| **Baseline Full-threshold Bipolar I or II Disorder** | Sleep Efficiency | 1.08 (0.99-1.19) | 0.121 | 0.724 | Age, Sex |
| **Baseline Full-threshold Psychotic Disorder** | Sleep Efficiency | 1.33 (1.11-1.66) | 0.005 | 0.03 | Age, Sex |

Results presented are logistic regression models adjusted for age and sex. Adjusted p values are Bonferroni corrected per outcome variable. CI = Confidence interval. MLE = Mania-like experiences. PLE = Psychosis-like experiences.

**Supplementary Table S2.** Prediction of specific longitudinal outcomes from sleep-wake measures

| Outcome | Predictors | Odds Ratio (95% CI) | P value | Adjusted p value | Covariates |
|---|---|---|---|---|---|
| **Full-threshold Bipolar I or II Disorder** | Sleep Duration | 1.83 (1.11-3.05) | 0.018 | 0.105 | Age, Sex |
| **Full-threshold Psychotic Disorder** | Sleep Duration | 2.93 (1.33-7.70) | 0.014 | 0.082 | Age, Sex |

Results presented are logistic regression models adjusted for age and sex. Adjusted p values are Bonferroni corrected per outcome variable. CI = Confidence interval. MLE = Mania-like experiences. PLE = Psychosis-like experiences. SOFAS = Social and Occupational Functioning Assessment Scale. NEET = Not in Education Employment or Training.

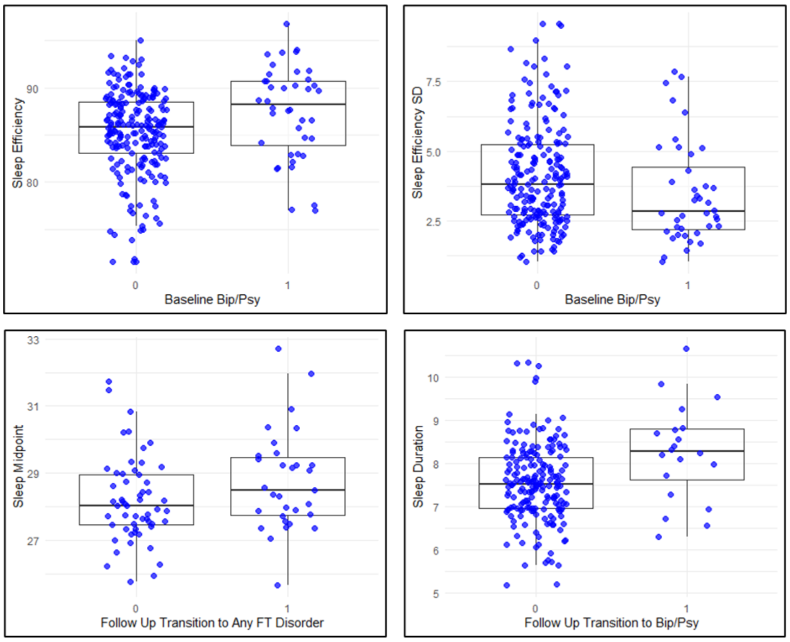


**Supplementary Figure S3.** Significant associations from logistic regression models for outcomes at baseline or across follow-up

**Supplementary Table S3.** Cox proportional hazards models predicting time to transition to specific follow-up outcomes from sleep wake measures

| Outcome | Predictors | Hazard Ratio (95% CI) | P value |
|---|---|---|---|
| **Full-threshold Bipolar I or II Disorder** | Age | 0.99 (0.87, 1.13) | 0.881 |
| | Sex (Male) | 0.29 (0.07, 1.31) | 0.107 |
| | Sleep Midpoint | 1.16 (0.77, 1.77) | 0.475 |
| | Sleep Duration | 1.54 (0.92, 2.60) | 0.101 |
| | Sleep Efficiency | 1.01 (0.88, 1.17) | 0.884 |
| | SD of Midpoint | 0.84 (0.21, 3.35) | 0.805 |
| | SD of Duration | 1.04 (0.28, 3.77) | 0.958 |
| | SD of Efficiency | 0.99 (0.68, 1.43) | 0.959 |
| **Full-threshold Psychotic Disorder** | Age | 1.04 (0.84, 1.29) | 0.729 |
| | Sex (Male) | 12.58 (2.05, 77.18) | 0.006 |
| | Sleep Midpoint | 0.86 (0.46, 1.62) | 0.638 |
| | Sleep Duration | 2.60 (1.07, 6.29) | 0.035 |
| | Sleep Efficiency | 1.02 (0.81, 1.29) | 0.856 |
| | SD of Midpoint | 0.13 (0.01, 2.66) | 0.184 |
| | SD of Duration | 4.02 (0.44, 36.33) | 0.216 |
| | SD of Efficiency | 1.27 (0.72, 2.27) | 0.408 |

CI = Confidence Interval. SD = Standard Deviation

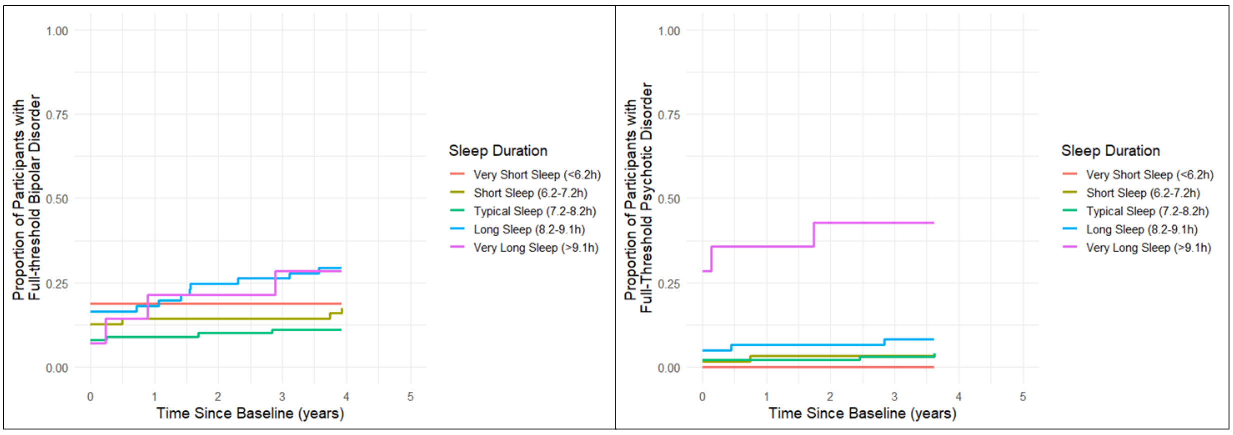


**Supplementary Figure S4.** Cumulative incidence of Full Threshold Bipolar or Psychotic Disorder by Sleep Duration Groups

The following analyses replicate analyses in tables 3, 4, and 5 in the main manuscript, while adding quadratic terms for sleep midpoint and sleep duration

**Supplementary Table S4.** Associations between sleep-wake measures and clinical characteristics at baseline

| Characteristic | Predictors | Term | Odds Ratio (95% CI) | P value |
|---|---|---|---|---|
| **Any Full-threshold Disorder** | Sleep Midpoint | Linear | 1.05 (0.85-1.29) | 0.680 |
| | | Quadratic | 0.99 (0.89-1.11) | 0.898 |
| | Sleep Duration | Linear | 1.19 (0.86-1.64) | 0.304 |
| | | Quadratic | 1.29 (1.01-1.66) | 0.043* |
| **Full-threshold Bipolar or Psychotic Disorder** | Sleep Midpoint | Linear | 0.98 (0.75-1.28) | 0.892 |
| | | Quadratic | 1.00 (0.88-1.14) | 0.997 |
| | Sleep Duration | Linear | 1.21 (0.85-1.72) | 0.288 |
| | | Quadratic | 1.14 (0.94-1.40) | 0.187 |
| **Other Full-threshold Disorder (Depressive or Anxious Disorders)** | Sleep Midpoint | Linear | 1.08 (0.88-1.31) | 0.463 |
| | | Quadratic | 1.01 (0.91-1.12) | 0.833 |
| | Sleep Duration | Linear | 1.02 (0.78-1.33) | 0.870 |
| | | Quadratic | 1.04 (0.88-1.23) | 0.612 |

Results presented are logistic regression models adjusted for age and sex. Adjusted p values are Bonferroni corrected per outcome variable. CI = Confidence interval. * p<.05

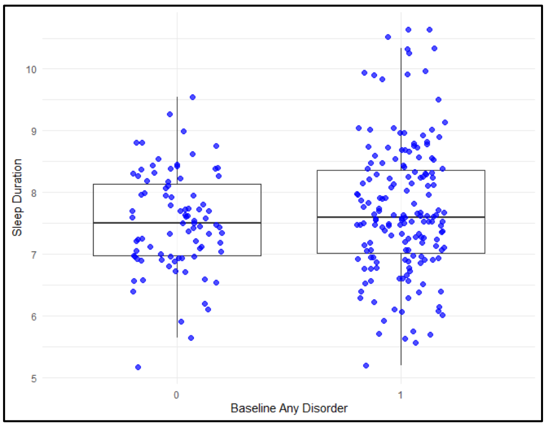


**Supplementary Figure S5** Significant quadratic association from logistic regression model between quadratic sleep duration and any full-threshold disorder at baseline

**Supplementary Table S5.** Associations between sleep-wake measures and clinical outcomes across longitudinal follow-up

| Outcome | Predictors | Term | Odds Ratio (95% CI) | P value |
|---|---|---|---|---|
| **Any Full-threshold Disorder** | Sleep Midpoint | Linear | 1.43 (0.96-2.13) | 0.075 |
| | | Quadratic | 1.02 (0.85-1.23) | 0.821 |
| | Sleep Duration | Linear | 1.22 (0.67-2.20) | 0.518 |
| | | Quadratic | 1.31 (0.82-2.11) | 0.261 |
| **Full-threshold Bipolar or Psychotic Disorder** | Sleep Midpoint | Linear | 1.03 (0.73-1.44) | 0.886 |
| | | Quadratic | 1.03 (0.87-1.22) | 0.746 |
| | Sleep Duration | Linear | 2.15 (1.12-4.11) | 0.021 |
| | | Quadratic | 1.03 (0.73-1.45) | 0.863 |
| **Other Full-threshold Disorder (Depressive or Anxious Disorders)** | Sleep Midpoint | Linear | 1.27 (0.84-1.92) | 0.261 |
| | | Quadratic | 1.04 (0.86-1.26) | 0.702 |
| | Sleep Duration | Linear | 0.95 (0.45-2.01) | 0.886 |
| | | Quadratic | 1.13 (0.63-2.01) | 0.684 |

Results presented are logistic regression models adjusted for age and sex. Adjusted p values are Bonferroni corrected per outcome variable. CI = Confidence interval.

**Table S6.** Cox proportional hazards models predicting time to transition to follow-up outcomes from sleep wake measures

| Outcome | Predictors | Hazard Ratio (95% CI) | P value |
|---|---|---|---|
| **Any Full-threshold Disorder** | Age | 1.00 (0.88, 1.13) | 0.954 |
| | Sex (Male) | 0.79 (0.26, 2.45) | 0.689 |
| | Sleep Midpoint (linear) | 0.96 (0.53, 1.73) | 0.883 |
| | Sleep Midpoint (quadratic) | 0.91 (0.79, 1.04) | 0.151 |
| | Sleep Duration (linear) | 1.68 (0.82, 3.45) | 0.159 |
| | Sleep Duration (quadratic) | 1.32 (0.87, 2.00) | 0.190 |
| | Sleep Efficiency | 1.01 (0.87, 1.16) | 0.906 |
| | SD of Midpoint | 0.42 (0.07, 2.53) | 0.343 |
| | SD of Duration | 3.32 (0.83, 13.22) | 0.089 |
| | SD of Efficiency | 1.21 (0.93, 1.57) | 0.155 |
| **Full-threshold Bipolar or Psychotic Disorder** | Age | 1.02 (0.89, 1.16) | 0.775 |
| | Sex (Male) | 1.93 (1.01, 3.70) | 0.046 |
| | Sleep Midpoint (linear) | 1.05 (0.79, 1.40) | 0.742 |
| | Sleep Midpoint (quadratic) | 0.94 (0.64, 1.39) | 0.772 |
| | Sleep Duration (linear) | 1.05 (0.90, 1.22) | 0.567 |
| | Sleep Duration (quadratic) | 1.05 (0.77, 1.44) | 0.756 |
| | Sleep Efficiency | 0.90 (0.27, 3.04) | 0.865 |
| | SD of Midpoint | 0.69 (0.21, 2.34) | 0.555 |
| | SD of Duration | 0.99 (0.89, 1.11) | 0.900 |
| | SD of Efficiency | 1.48 (0.55, 4.00) | 0.434 |
| **Other Full-threshold Disorder (Depressive or Anxious Disorders)** | Age | 1.03 (0.94, 1.14) | 0.512 |
| | Sex (Male) | 0.92 (0.36, 2.33) | 0.861 |
| | Sleep Midpoint (linear) | 1.08 (0.72, 1.60) | 0.719 |
| | Sleep Midpoint (quadratic) | 0.96 (0.82, 1.12) | 0.586 |
| | Sleep Duration (linear) | 0.59 (0.31, 1.09) | 0.094 |
| | Sleep Duration (quadratic) | 0.93 (0.57, 1.51) | 0.766 |
| | Sleep Efficiency | 1.08 (0.92, 1.25) | 0.349 |
| | SD of Midpoint | 0.21 (0.04, 1.07) | 0.06 |
| | SD of Duration | 2.78 (0.76, 10.20) | 0.124 |
| | SD of Efficiency | 1.09 (0.81, 1.47) | 0.561 |

CI = Confidence interval. SD = Standard Deviation.
